# ROR1 is a novel putative druggable target for diffuse glioma

**DOI:** 10.1101/2021.12.03.21267166

**Authors:** Miya John, Padmashree Rao, Humaira Noor, Caroline Ford

## Abstract

The cell surface receptor ROR1 is a therapeutic target of growing interest in oncology; however, its role in glioma has not been established thus far. This study analyzed associations between ROR1 mRNA expression and clinical outcomes, and histological and molecular subtypes in four independent glioma (grades II-IV) transcriptomic datasets (The Cancer Genome Atlas-GBMLGG, Chinese Glioma Genome Atlas, Repository for Molecular Brain Neoplasia, and GSE16011), encompassing a total of 2,388 cases. The data strongly suggests that ROR1 may be associated with poorer outcomes and more aggressive disease. Taken together, ROR1 should be further examined as a novel putative druggable target for glioma, a cancer that currently has very limited therapeutic options.

## Introduction

Gliomas account for ∼80% of primary brain cancer (1), and have four histological grades (grade I to IV), with grade I primarily occurring in pediatric cases and higher grades in adults. Adult diffuse low-grade (World Health Organization [WHO] grade II) and intermediate-grade (WHO grade III) gliomas are together classified as lower-grade gliomas (LGG) (2). The histologic subtypes of LGG include astrocytoma, oligodendroglioma and mixed gliomas. Glioblastoma (GBM) are grade IV tumors that can arise *de novo* (primary) or progress from LGGs (secondary). LGG and GBM are highly infiltrative with a propensity to develop resistance to standard chemotherapy. With limited treatment options, LGG and GBM are mostly fatal with survival ranging from months to 15 years.

Cancer stem cells (CSCs) are a small sub-population of tumor cells that are immortal and with the capacity to differentiate into heterogeneous tumor cell types, are the precursors for *de novo* tumor formation (3). In gliomas like other cancers, CSCs are chemotherapy and radiation-resistant, as well as more infiltrative and metastatic (3). A growing body of evidence suggests that CSCs in adult gliomas seeding more aggressive and invasive tumor growth following current therapeutic strategies including surgical debulking, radiotherapy and chemotherapy underlies treatment failure, making them attractive therapeutic targets (3).

Receptor Tyrosine Kinase Like Orphan Receptor 1 (ROR1) is a cell-surface receptor associated with stemness and poor clinical outcomes in various solid and hematological cancers (4-7). ROR1 is primarily expressed during embryonic development and considered mostly turned off in adult tissues (8). ROR1 targeted therapeutics for oncology has recently received significant attention with numerous preclinical studies and Phase 1/2 clinical trials under way (4-8); however, its potential in treating glioma has not been previously explored.

Advances in next generation sequencing technologies have resulted in important breakthroughs related to molecular classification and clinical decision making for glioma. Molecular markers associated with favorable prognosis of glioma include inactivating mutations in the isocitrate dehydrogenase (IDH) 1/2 genes and codeletions (codel) of chromosome arms 1p and 19q (2). Multi-omic projects including The Cancer Genome Atlas (TCGA), Chinese Glioma Genome Atlas (CGGA) and Repository for Molecular Brain Neoplasia (REMBRANDT) among others have provided a treasure-trove of data (2, 9-11); however, a substantial improvement in therapeutic outcomes remains to be realized. The discovery of novel associations between genes and clinical outcomes could uncover promising new therapies for glioma. This study examines the association between ROR1 mRNA expression and clinical outcomes in glioma, as well as its histological and molecular subtypes to find a consistent association between high ROR1 expression and poor prognosis in glioma, identifying ROR1 as a novel putative therapeutic target for glioma.

## Results

To examine a possible association between ROR1 and clinical outcomes in glioma, meta-analysis of four independent glioma transcriptomic datasets was performed (Table 1). Only cases with reported histology of astrocytoma, oligodendroglioma, mixed glioma (or oligoastrocytoma) or GBM, and with ROR1 mRNA expression data were included in the analysis, which included a total of 2,388 cases (Table 1). The LGG cohorts included astrocytoma, oligodendroglioma and mixed glioma cases. Kaplan-Meier survival analysis identified a strong association between lower ROR1 mRNA expression and higher overall survival in the LGG cohorts of TCGA, CGGA, REMBRANDT and GSE16011 datasets (Fig 1A-D). Consistently, a similar effect was seen in the astrocytoma cohorts of TCGA, CGGA and REMBRANDT datasets (Fig 1E-G). Although not significant (p = 0.085), a similar trend was seen with the relatively smaller (n=29) astrocytoma cohort of GSE16011 (Fig 1H). Lower ROR1 expression also significantly correlated with higher overall survival in the oligodendroglioma cohort of TCGA (Fig 1I), while a similar trend with p values of 0.18, 0.2 and 0.13 were observed for the CGGA, REMBRANDT and GSE16011 datasets respectively (Fig 1J-L). In GBM, ROR1 expression was not associated with differential survival in the TCGA and REMBRANDT datasets (Fig 1M and O); however, in the CGGA and GSE16011 datasets, lower ROR1 expression significantly associated with higher overall survival (Fig 1N and P). Amongst 41 TCGA datasets, Cox multivariate regression analysis found the strongest association of ROR1 mRNA expression with overall survival, adjusted by age, gender race and tumor stage in LGG (Fig 1Q and SI Appendix 2).

**Table 1.** Clinical characteristics of glioma datasets.

Clinical characteristics of lower-grade glioma cohorts
| Dataset | TCGA-GBMLGG | CGGA | REMBRANDT | GSE16011 |
| --- | --- | --- | --- | --- |
| **Characteristic** |  |  |  |  |
| **Histologic type and grade - n/total n (%)** |  |  |  |  |
| Astrocytoma | 194/667 (29.1) | 387/1009 (38.4) | 147/444 (33.1) | 29/268 (10.8) |
| Grade II | 63/194 (32.5) | 173/387 (44.7) | 64/147 (43.5) | 13/29 (44.8) |
| Grade III | 131/194 (67.5) | 214/387 (55.3) | 58/147 (39.5) | 16/29 (55.2) |
| Unreported grade | 0 | 0 | 25/147 (17) | 0 |
| Oligodendroglioma | 191/667 (28.6) | 206/1009 (20.4) | 67/444 (15.1) | 52/268 (19.4) |
| Grade II | 110/191 (57.6) | 108/206 (52.4) | 30/67 (44.8) | 8/52 (15.4) |
| Grade III | 79/191 (41.4) | 98/206 (47.6) | 23/67 (34.3) | 44/52 (84.6) |
| Unreported grade | 2/191 (1.0) | 0 | 14/67 (20.9) | 0 |
| Mixed glioma | 130/667 (19.5) | 30/1009 (3.0) | 11/444 (2.5) | 28/268 (10.4) |
| Grade II | 73/130 (56.2) | 9/30 (30.0) | 4/11 (36.4) | 3/28 (10.7) |
| Grade III | 56/130 (43.1) | 21/30 (70.0) | 3/11 (27.3) | 25/28 (89.3) |
| Unreported grade | 1/130 (0.8) | 0 | 4/11 (36.4) | 0 |
| GBM | 152/667 (22.8) | 386/1009 (38.3) | 219/444 (49.3) | 159/268 (59.3) |
| Grade IV | 149/152 (98.0) | 386/386 (100) | 130/219 (59.4) | 159/159 (100) |
| Unreported grade | 3/152 (2.0) | 0 | 87/219 (39.7) | 0 |
| Other grades | 0 | 0 | 2/219 (0.9) | 0 |
| **Histologic type and IDH status - n/total n (%)** |  |  |  |  |
| Astrocytoma |  |  |  |  |
| IDH1/2 wt | 57/194 (29.4) | 129/387 (33.3) | - | 16/29 (55.2)* |
| IDH1/2 mt | 135/194 (69.6) | 246/387 (63.6) | - | 9/29 (31.0) * |
| Not reported | 2/194 (1.0) | 12/387 (3.1) |  | 4/29 (13.8) * |
| Oligodendroglioma |  |  |  |  |
| IDH1/2 wt | 21/191 (11.0) | 14/206 (6.8) | - | 17/52 (32.7) * |
| IDH1/2 mt | 169/191 (88.5) | 180/206 (87.4) | - | 27/52 (51.9) * |
| Not reported | 1/191 (0.5) | 12/206 (5.8) |  | 8/52 (15.4) * |
| Mixed glioma |  |  |  |  |
| IDH1/2 wt | 16/130 (12.3) | 0/30 (0) | - | 8/28 (28.6) * |
| IDH1/2 mt | 114/130 (87.7) | 12/30 (40.0) | - | 11/28 (39.3) * |
| Not reported | 0/130 (0) | 18/30 (60.0) |  | 9/28 (32.1) * |
| GBM |  |  |  |  |
| IDH1/2 wt | 138/152 (90.8) | 287/386 (74.4) | - | 95/159 (59.7) |
| IDH1/2 mt | 10/152 (6.6) | 89/386 (23.1) | - | 33/159 (20.8) |
| Not reported | 4/152 (2.6) | 10/386 (2.6) |  | 31/159 (19.5) |
| **Age at diagnosis – years** |  |  |  |  |
| Median ± SD | 47 ± 15.27 | 42.5 ± 12.27 | 50 to 54 ± 14.87 | 51.9 ± 14.17 |
| Range | 14 – 89 | 8 - 79 | 15 to 19 – 85 to 89 | 14 - 81 |
| unreported | 58 | 1 | 46 | 0 |
| **Gender** |  |  |  |  |
| Male | 355/667 (53.2) | 595/1009 (59.0) | 214/444 (48.2) | 180/268 (67.2) |
| Female | 254/667 (38.1) | 414/1009 (41.0) | 126/444 (28.4) | 88/268 (32.8) |
| Unreported | 58/667 (8.7) |  | 104/444 (23.4) | 0 |
| **Recurrence** |  |  |  |  |
| Primary | 616/667 (92.4) | 651/1009 (64.5) |  |  |
| Recurrent | - | 330/1009 (32.7) |  |  |
| Secondary | - | 28/1009 (2.8) |  |  |
| Unreported | 51/667 (7.6) | - |  |  |

\* Only IDH1 tested.

**Figure 1.**
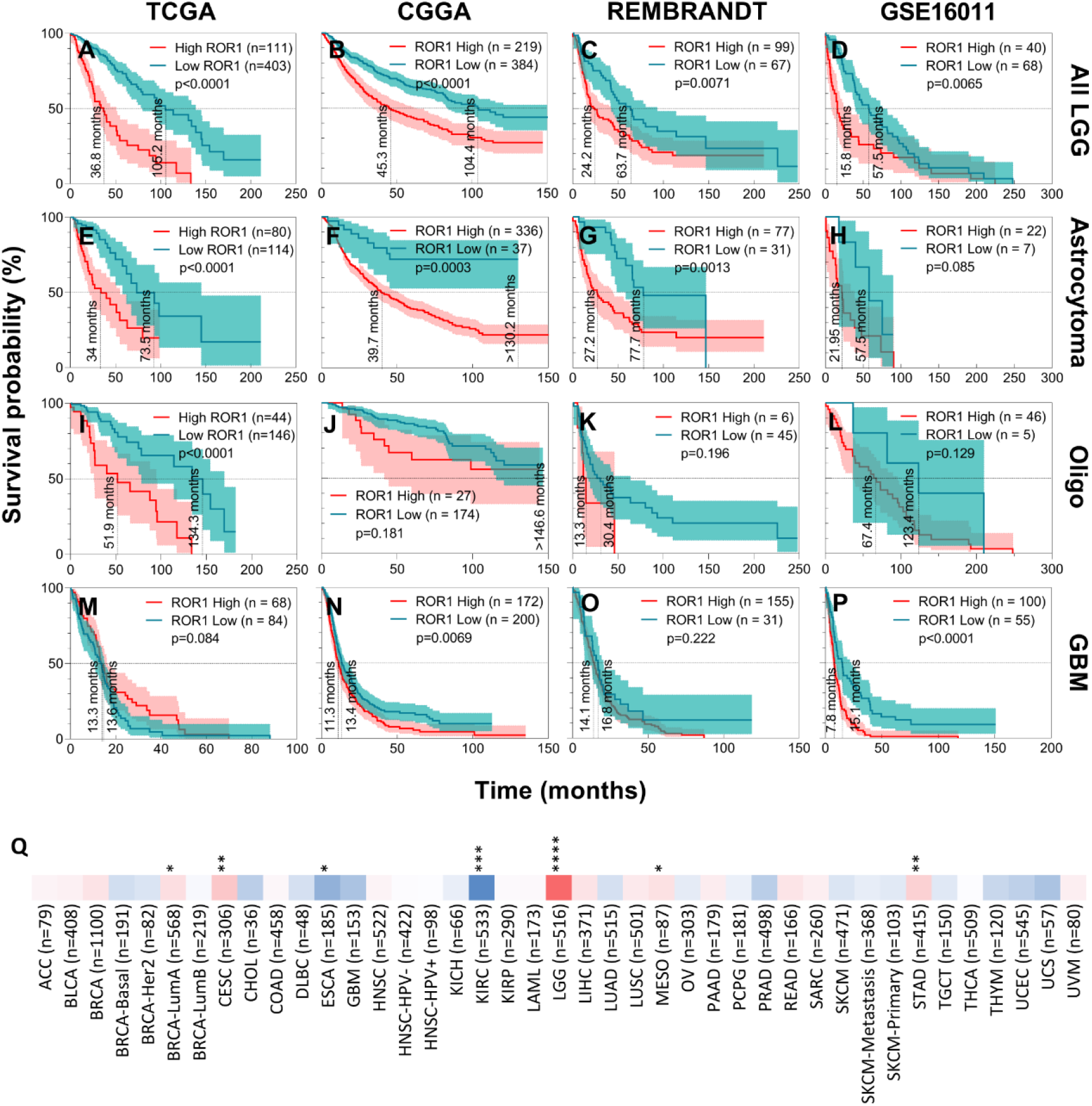
Association between ROR1 and clinical outcomes in Glioma. Kaplan-Meier survival analysis of correlation between ROR1 mRNA expression in glioma tumor samples and overall survival using the Log-rank (Mantel-Cox) test in all LGG (A-D), astrocytoma (E-H), oligo (oligodendroglioma; I-L) and GBM (M-P) cohorts of the TCGA, CGGA, REMBRANDT and GSE16011 datasets (left to right columns respectively). Red, high expression; blue, low expression. (Q) Cox proportional hazard model of the effect of ROR1 mRNA expression on overall survival, adjusted by age, gender, race and tumor stage in TCGA datasets. (* p<0.05, ** p<0.01, *** p<0.001, **** p<0.0001)

Grade IV and GBM tumors had highest ROR1 mRNA expression compared to less aggressive histologies (Fig 2A, B). Amongst the three datasets that included data for IDH1/2 mutation and 1p/19q codel, IDH mutated tumors had significantly lower ROR1 mRNA expression in the LGG cohorts of TCGA and CGGA datasets and in the GBM cohort of the CGGA dataset (Fig 2C). Furthermore, tumors with 1p/19q codel had significantly lower ROR1 mRNA expression in the LGG cohorts of all three datasets and in the GBM cohort of the CGGA dataset (Fig 2D).

**Figure 2.**
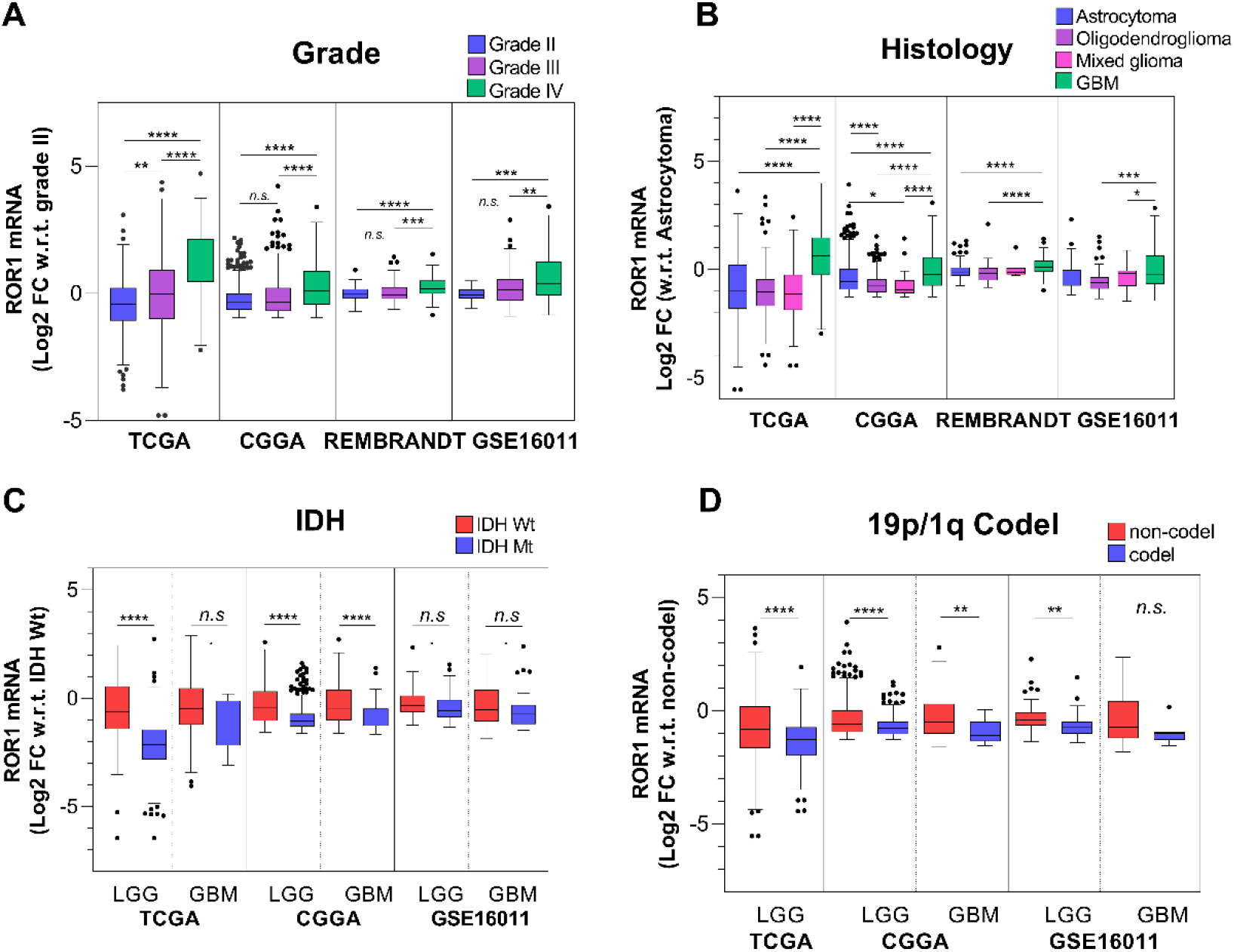
ROR1 mRNA expression in histological and molecular subtypes of glioma. ROR1 mRNA expression in grade II, III and IV tumors (A), astrocytoma, oligodendroglioma, mixed glioma and GBM histological subtypes (B), IDH wildtype and mutant tumors of LGG and GBM cohorts (C), and tumors with 1p/19q non-codel and codel in LGG and GBM cohorts (D) of TCGA, CGGA, REMBRANDT and GSE16011 datasets. Statistical tests employed are detailed in methods; *p<0.05, **p<0.01, ***p<0.001, ****p<0.0001.

## Discussion

Meta-analysis of four independent multi-omic datasets including survival data for 2,388 LGG and GBM patients identified strong associations between high ROR1 mRNA expression in the tumor and poor overall survival of LGG patients (Fig 1A-D). Consistently, as shown in Fig 1E-L, a similar trend was observed in both astrocytoma and oligodendroglioma cohorts, albeit significant in only three of four astrocytoma cohorts (TCGA, CGGA and REMBRANDT) and one of four oligodendroglioma cohorts (TCGA). It should be noted that smaller sample size of the astrocytoma cohort within the GSE16011 dataset and the oligodendroglioma cohorts within the CGGA, REMBRANDT and GSE16011 is a contributing factor for reduced significance of the association. High ROR1 mRNA expression in the tumor was associated with poor overall survival in two of four GBM cohorts (Fig 1N and P). Due to the homogenously low survival time for GBM patients, the power to test the effect of a gene on survival is very low, requiring large cohort sizes. The lack of an association in the TCGA and REMBRANDT cohorts (Fig 1M and O) does not necessarily mean that ROR1 did not influence survival outcomes, rather that sample sizes may not have been large enough to detect an effect. This is evidenced by a significant (p=0.038) association between high ROR1 mRNA expression and poor survival in a larger (n=488) TCGA study of GBM alone (data not shown). Interestingly, the role of ROR1 has now been well studied in multiple cancers excluding LGG and it was in this cancer that the strongest association between ROR1 mRNA expression and overall survival, adjusted by age, gender race and tumor stage was found by Cox multi-variate analysis (Fig 1Q).

ROR1 mRNA expression consistently increased with increasing tumor grades - an ANOVA test for linear trend from grade II to IV was statistically significant (p<0.0001) in all four datasets and ROR1 expression in grade IV was consistently higher than grade II and III (Fig 2A). However, levels in grade III compared to grade II was not consistently higher (Fig 2A), possibly due to the prevalence of intra- and inter-observer variability between grade II and III tumors (12). Between histological sub-types, ROR1 was most highly expressed in GBM (Fig 2B). Furthermore, ROR1 mRNA expression was higher in the more aggressive IDH wt tumors within the LGG and GBM cohorts of the CGGA dataset and in the LGG cohort of the TCGA dataset (Fig 2C). More consistently, gliomas with 1p/19q codel, a favorable prognostic biomarker, had lower ROR1 mRNA expression (Fig 2D). While no studies have thus far examined the association between ROR1 expression and survival in glioma, knockdown of ROR1 in GBM cell lines has been reported to reduce stemness, invasion and metastasis (7). Furthermore, an association between higher WNT5A (an activating ligand for ROR1) and poor overall survival in glioma patients has been previously reported (13). Taken together, the inference from the association between lower ROR1 levels and better survival as well as less aggressive disease in glioma is that pharmacological inhibition of ROR1 may improve survival. This is an important finding because targeting ROR1 may be safe for clinical translation as well as effective against the CSC population of the tumor. Furthermore, inhibition of ROR1 may inhibit infiltrative cells that cannot be surgically resected.

ROR1 as a therapeutic target for glioma is not without limitations. Firstly, antibody-based therapies being developed may not cross the blood brain barrier. However, the monoclonal antibody cirmtuzumab has a long half-life of 32.4 days (14), and treatment during surgery may offer some benefit. Furthermore, alternative approaches more suitable for brain tumors are also currently under investigation. Secondly, there is significant heterogeneity within the CSC population (3), and although this study identifies one putative CSC driver of interest, ideally, multiple CSC drivers may need to be concurrently inhibited for substantial therapeutic benefit. Preclinical *in vitro* and *in vivo* models can further examine the role of ROR1 in glioma tumorigenesis to establish if its inhibition can reduce tumor recurrence and improve survival. The ROR1 transcript variant ENST00000371079 has been deemed the principal variant of ROR1; however, we have previously demonstrated that ENST00000545203 which lacks a signal peptide for cell surface localisation may be the predominantly expressed transcript variant in healthy tissues and cancer (16). Hence, the sub-cellular localization and function of ROR1 needs to be assessed before therapeutics can be designed.

In conclusion, this study represents the first report of an association between clinical outcomes in glioma and ROR1, a known CSC marker that is druggable and hence identifies ROR1 as a novel putative therapeutic target for glioma that can be rapidly translated to the clinic.

## Methods

*Patients:* ROR1 tumor mRNA expression and associated clinical data from TCGA-GBMLGG, CGGA, REMBRANDT and GSE16011 were downloaded from GlioVis portal (15). Methods are detailed in *SI Appendix 1*.

## Data Availability

All data produced in the present study are available upon reasonable request to the authors.

## Acknowledgments

We gratefully acknowledge the individuals and institutions that have provided samples and data analyzed in this study. We gratefully acknowledge Professor John R. Adler Jr. and Professor Alexander Muacevic for reviewing the manuscript and providing important feedback.

## Notes

### Competing Interest Statement

The authors have declared no competing interest.

### Funding Statement

This project was supported in part by a Major Pilot Grant to CF from the Translational Cancer Research Group, an initiative of the Cancer Institute New South Wales, and an Early Career Research Grant to MJ from Tour de Cure.

### Author Declarations

This study involves only openly available human data, which can be obtained from: http://gliovis.bioinfo.cnio.es/

