## Supplementary Information Appendix 1 for "ROR1 is a novel putative druggable target for diffuse glioma"

**Methods**

*Patients:* ROR1 tumor mRNA expression and associated clinical data from glioma datasets were downloaded from GlioVis portal (http://gliovis.bioinfo.cnio.es/) (TCGA-LGG on 18/05/2020, CGGA on 28/10/2020, REMBRANDT on 3/02/2021, GSE16011 on 5/02/2021, TCGA-GBMLGG on 18/09/2021). Only cases with histology reported as astrocytoma, oligodendroglioma, mixed glioma (or oligoastrocytoma) or GBM were included and samples without ROR1 mRNA expression data were excluded. In the TCGA-GBMLGG dataset, clinical and molecular data for 669 glioma patients was downloaded from GlioVis portal and incomplete clinical data was queried on the cBioPortal. Two cases were excluded from all further analysis due to lack of histological data, leaving a cohort of 667 cases. For the CGGA dataset, clinical and molecular data for 1018 glioma patients was downloaded. Five and four cases were excluded due to missing histology and RNA expression data respectively, resulting in a cohort of 1009. For the REMBRANDT dataset, clinical and molecular data for 580 glioma patients was downloaded from which 444 cases were included for further based on histology. For the GSE16011 dataset, clinical and molecular data for 284 glioma patients was downloaded from which 268 cases were included for further based on histology.

*Survival analysis*: Association between ROR1 mRNA expression and overall survival was estimated with Kaplan-Meier survival curves within the whole LGG, astrocytoma, oligodendroglioma and GBM cohorts of each dataset. Optimal cut-off values (based on maximally selected rank statistics) were determined with the surv_cutpoint function of the survminer R package. Kaplan-Meier survival curves were plotted on GraphPad Prism 8 software after dividing each cohort into high and low ROR1 expression groups based on the optimal cut-off values. Log-rank (Mantel-Cox) test was employed to compare curves between groups. Number of patients, 95% confidence intervals (CI) and median survival in each group, and p values are reported in the figure. $P < 0.05$ was considered statistically significant. Cox proportional hazard model of the effect of ROR1 mRNA expression on overall survival, adjusted by age, gender, race and tumor stage in TCGA datasets was analyzed on TIMER 2.0. Normalized coefficients of ROR1 mRNA expression in the Cox model was represented in a heatmap along with statistical significance.

*Analysis of ROR1 mRNA expression in histological and molecular subtypes of glioma*: Ordinary one-way ANOVA test or unpaired t-test were employed to examine statistically significant difference between 3≥ or 2 groups respectively.
