## Supplementary Information Appendix 2 for "ROR1 is a novel putative druggable target for diffuse glioma"

**TCGA Abbreviations:**
ACC (adrenocortical carcinoma), BLCA (bladder urothelial carcinoma), BRCA (breast invasive carcinoma), CESC (cervical squamous cell carcinoma and endocervical adenocarcinoma), CHOL (cholangiocarcinoma), COAD (colon adenocarcinoma), DLBC (lymphoid neoplasm diffuse large B-cell lymphoma), ESCA (esophageal carcinoma), GBM (glioblastoma multiforme), HNSC (head and neck squamous cell carcinoma), KICH (kidney chromophobe), KIRC (kidney renal clear cell carcinoma), KIRP (kidney renal papillary cell carcinoma), LAML (acute myeloid leukemia), LGG (brain lower grade glioma), LIHC (liver hepatocellular carcinoma), LUAD (lung adenocarcinoma), LUSC (lung squamous cell carcinoma), MESO (mesothelioma), OV (ovarian serous cystadenocarcinoma), PAAD (pancreatic adenocarcinoma), PCPG (pheochromocytoma and paraganglioma), PRAD (prostate adenocarcinoma), READ (rectum adenocarcinoma), SARC (sarcoma), SKCM (skin cutaneous melanoma), STAD (stomach adenocarcinoma), TGCT (testicular germ cell tumours), THCA (thyroid carcinoma), THYM (thymoma), UCEC (uterine corpus endometrial carcinoma), UCS (uterine carcinosarcoma) and UVM (uveal melanoma).

**Multivariate Analysis**
**ROR1 in ACC (n=79):**
Model: Surv(OS, EVENT) ~ `ROR1` + Age + Gender + Race
68 patients with 26 dying ( 11 missing obs. )

| | coef | HR | se(coef) | 95%CI_l | 95%CI_u | z | p | signif |
|---|---|---|---|---|---|---|---|---|
| ROR1 | 0.542 | 1.72 | 0.454 | 0.706 | 4.189 | 1.194 | 0.233 | |
| Age | 0.005 | 1.005 | 0.014 | 0.978 | 1.032 | 0.342 | 0.732 | |
| Gendermale | 0.055 | 1.056 | 0.401 | 0.481 | 2.318 | 0.136 | 0.891 | |
| RaceBlack | 1.421 | 4.143 | 11699.01 | 0 | Inf | 0 | 1 | <NA> |
| RaceWhite | 18.409 | 98814492.01 | 10046.1 | 0 | Inf | 0.002 | 0.999 | |

Rsquare = 0.051 (max possible = 9.43e-01 )
Likelihood ratio test p = 6.17e-01
Wald test p = 9.16e-01
Score (logrank) test p = 7.44e-01

**ROR1 in BLCA (n=408):**
Model: Surv(OS, EVENT) ~ `ROR1` + Age + Gender + Race + Stage
385 patients with 169 dying ( 23 missing obs. )

| | coef | HR | se(coef) | 95%CI_l | 95%CI_u | z | p | signif |
|---|---|---|---|---|---|---|---|---|
| ROR1 | 0.196 | 1.216 | 0.146 | 0.913 | 1.62 | 1.338 | 0.181 | |
| Age | 0.033 | 1.033 | 0.008 | 1.017 | 1.05 | 3.962 | 0 | *** |
| Gendermale | -0.156 | 0.855 | 0.173 | 0.609 | 1.201 | -0.903 | 0.367 | |
| RaceBlack | 0.509 | 1.664 | 0.438 | 0.706 | 3.923 | 1.163 | 0.245 | |
| RaceWhite | 0.036 | 1.037 | 0.354 | 0.518 | 2.076 | 0.102 | 0.919 | |
| Stage2 | 14.467 | 1918663.287 | 1918.804 | 0 | Inf | 0.008 | 0.994 | |
| Stage3 | 14.872 | 2875091.382 | 1918.804 | 0 | Inf | 0.008 | 0.994 | |
| Stage4 | 15.423 | 4990663.309 | 1918.804 | 0 | Inf | 0.008 | 0.994 | |

Rsquare = 0.122 (max possible = 9.9e-01 )
Likelihood ratio test p = 4.19e-08
Wald test p = 2.13e-07
Score (logrank) test p = 5.67e-08

**ROR1 in BRCA (n=1100):**
Model: Surv(OS, EVENT) ~ `ROR1` + Age + Gender + Race + Stage
976 patients with 136 dying ( 124 missing obs. )

| | coef | HR | se(coef) | 95%CI_l | 95%CI_u | z | p | signif |
|---|---|---|---|---|---|---|---|---|

| | | | | | | | | |
|---|---|---|---|---|---|---|---|---|
| ROR1 | 0.191 | 1.211 | 0.103 | 0.989 | 1.482 | 1.855 | 0.064 | . |
| Age | 0.035 | 1.036 | 0.007 | 1.022 | 1.05 | 5.018 | 0 | *** |
| Gendermale | -0.516 | 0.597 | 1.012 | 0.082 | 4.337 | -0.51 | 0.61 | |
| RaceBlack | 0.126 | 1.134 | 0.616 | 0.339 | 3.789 | 0.204 | 0.838 | |
| RaceWhite | -0.176 | 0.838 | 0.595 | 0.261 | 2.69 | -0.297 | 0.767 | |
| Stage2 | 0.579 | 1.785 | 0.281 | 1.029 | 3.096 | 2.062 | 0.039 | * |
| Stage3 | 1.244 | 3.471 | 0.296 | 1.942 | 6.205 | 4.198 | 0 | *** |
| Stage4 | 2.697 | 14.834 | 0.38 | 7.045 | 31.233 | 7.099 | 0 | *** |

Rsquare = 0.075 (max possible = 7.93e-01 )
Likelihood ratio test p = 3.43e-13
Wald test p = 6.74e-17
Score (logrank) test p = 6.65e-23

**ROR1 in BRCA-Basal (n=191):**
Model: Surv(OS, EVENT) ~ `ROR1` + Age + Race + Stage
177 patients with 26 dying ( 14 missing obs. )

| | coef | HR | se(coef) | 95%CI_l | 95%CI_u | z | p | signif |
|---|---|---|---|---|---|---|---|---|
| ROR1 | -0.085 | 9.18E-01 | 0.208 | 0.61 | 1.382 | -0.409 | 0.683 | |
| Age | 0.007 | 1.01E+00 | 0.017 | 0.975 | 1.041 | 0.437 | 0.662 | |
| RaceBlack | -1.026 | 3.58E-01 | 1.084 | 0.043 | 3.001 | -0.946 | 0.344 | |
| RaceWhite | -1.537 | 2.15E-01 | 1.124 | 0.024 | 1.946 | -1.368 | 0.171 | |
| Stage2 | 18.845 | 1.53E+08 | 6162.592 | 0 | Inf | 0.003 | 0.998 | |
| Stage3 | 20.225 | 6.08E+08 | 6162.592 | 0 | Inf | 0.003 | 0.997 | |
| Stage4 | 21.167 | 1.56E+09 | 6162.592 | 0 | Inf | 0.003 | 0.997 | |

Rsquare = 0.153 (max possible = 7.23e-01 )
Likelihood ratio test p = 1.22e-04
Wald test p = 3.22e-03
Score (logrank) test p = 8.56e-07

**ROR1 in BRCA-Her2 (n=82):**
Model: Surv(OS, EVENT) ~ `ROR1` + Age + Race + Stage
66 patients with 13 dying ( 16 missing obs. )

| | coef | HR | se(coef) | 95%CI_l | 95%CI_u | z | p | signif |
|---|---|---|---|---|---|---|---|---|
| ROR1 | -0.113 | 0.893 | 0.667 | 0.242 | 3.3 | -0.17 | 0.865 | |
| Age | 0.048 | 1.049 | 0.03 | 0.989 | 1.113 | 1.59 | 0.112 | |
| RaceBlack | -2.902 | 0.055 | 1.745 | 0.002 | 1.679 | -1.663 | 0.096 | . |
| RaceWhite | -1.511 | 0.221 | 1.438 | 0.013 | 3.692 | -1.051 | 0.293 | |
| Stage2 | -0.431 | 0.65 | 1.154 | 0.068 | 6.242 | -0.374 | 0.709 | |
| Stage3 | 0.674 | 1.961 | 1.204 | 0.185 | 20.769 | 0.559 | 0.576 | |
| Stage4 | NA | NA | 0 | NA | NA | NA | NA | <NA> |

Rsquare = 0.326 (max possible = 7.08e-01 )
Likelihood ratio test p = 2.22e-04
Wald test p = 1.14e-01
Score (logrank) test p = 6.28e-17

**ROR1 in BRCA-LumA (n=568):**
Model: Surv(OS, EVENT) ~ `ROR1` + Age + Gender + Race + Stage
513 patients with 59 dying ( 55 missing obs. )

| | coef | HR | se(coef) | 95%CI_l | 95%CI_u | z | p | signif |
|---|---|---|---|---|---|---|---|---|
| ROR1 | 0.355 | 1.426 | 0.17 | 1.022 | 1.991 | 2.088 | 0.037 | * |
| Age | 0.047 | 1.048 | 0.011 | 1.026 | 1.071 | 4.342 | 0 | *** |
| Gendermale | -15.492 | 0 | 3837.774 | 0 | Inf | -0.004 | 0.997 | |
| RaceBlack | -0.246 | 0.782 | 1.173 | 0.078 | 7.793 | -0.21 | 0.834 | |
| RaceWhite | 0.222 | 1.248 | 1.028 | 0.166 | 9.365 | 0.215 | 0.829 | |
| Stage2 | 0.458 | 1.581 | 0.363 | 0.776 | 3.224 | 1.261 | 0.207 | |
| Stage3 | 0.873 | 2.394 | 0.393 | 1.108 | 5.174 | 2.22 | 0.026 | * |
| Stage4 | 2.411 | 11.149 | 0.592 | 3.494 | 35.571 | 4.074 | 0 | *** |

Rsquare = 0.069 (max possible = 6.74e-01 )
Likelihood ratio test p = 1.35e-05
Wald test p = 1.13e-06
Score (logrank) test p = 1.29e-08

**ROR1 in BRCA-LumB (n=219):**
Model: Surv(OS, EVENT) ~ `ROR1` + Age + Gender + Race + Stage
181 patients with 31 dying ( 38 missing obs. )

| | coef | HR | se(coef) | 95%CI_l | 95%CI_u | z | p | signif |
|---|---|---|---|---|---|---|---|---|
| ROR1 | 0.227 | 1.254 | 0.339 | 0.645 | 2.439 | 0.668 | 0.504 | |
| Age | 0.034 | 1.035 | 0.016 | 1.002 | 1.069 | 2.068 | 0.039 | * |
| Gendermale | -0.337 | 0.714 | 1.069 | 0.088 | 5.803 | -0.316 | 0.752 | |
| RaceBlack | 16.305 | 12051610.07 | 4995.883 | 0 | Inf | 0.003 | 0.997 | |
| RaceWhite | 15.822 | 7439054.81 | 4995.883 | 0 | Inf | 0.003 | 0.997 | |
| Stage2 | 1.178 | 3.247 | 0.801 | 0.675 | 15.609 | 1.47 | 0.142 | |
| Stage3 | 1.773 | 5.888 | 0.819 | 1.182 | 29.339 | 2.164 | 0.03 | * |
| Stage4 | 2.557 | 12.9 | 0.963 | 1.952 | 85.239 | 2.654 | 0.008 | ** |

Rsquare = 0.081 (max possible = 7.35e-01 )
Likelihood ratio test p = 5.29e-02
Wald test p = 8.47e-02
Score (logrank) test p = 3.43e-02

**ROR1 in CESC (n=306):**
Model: Surv(OS, EVENT) ~ `ROR1` + Age + Race
255 patients with 63 dying ( 51 missing obs. )

| | coef | HR | se(coef) | 95%CI_l | 95%CI_u | z | p | signif |
|---|---|---|---|---|---|---|---|---|
| ROR1 | 0.69 | 1.993 | 0.224 | 1.284 | 3.092 | 3.077 | 0.002 | ** |
| Age | 0.02 | 1.02 | 0.009 | 1.001 | 1.039 | 2.096 | 0.036 | * |
| RaceBlack | 0.54 | 1.716 | 0.801 | 0.357 | 8.243 | 0.674 | 0.5 | |
| RaceWhite | 0.525 | 1.69 | 0.725 | 0.408 | 7.004 | 0.724 | 0.469 | |

Rsquare = 0.047 (max possible = 9.02e-01 )
Likelihood ratio test p = 1.61e-02
Wald test p = 7.51e-03
Score (logrank) test p = 6.62e-03

**ROR1 in CHOL (n=36):**
Model: Surv(OS, EVENT) ~ `ROR1` + Age + Gender + Race + Stage
36 patients with 18 dying ( 0 missing obs. )

| | coef | HR | se(coef) | 95%CI_l | 95%CI_u | z | p | signif |
|---|---|---|---|---|---|---|---|---|
| ROR1 | -0.298 | 0.742 | 0.28 | 0.429 | 1.285 | -1.064 | 0.287 | |
| Age | 0.017 | 1.017 | 0.023 | 0.972 | 1.064 | 0.733 | 0.464 | |
| Gendermale | 0.172 | 1.188 | 0.557 | 0.399 | 3.537 | 0.31 | 0.757 | |
| RaceBlack | 0.52 | 1.683 | 1.506 | 0.088 | 32.23 | 0.345 | 0.73 | |
| RaceWhite | -0.583 | 0.558 | 0.936 | 0.089 | 3.491 | -0.624 | 0.533 | |
| Stage2 | 0.892 | 2.44 | 0.697 | 0.622 | 9.572 | 1.279 | 0.201 | |
| Stage3 | -17.197 | 0 | 6920.755 | 0 | Inf | -0.002 | 0.998 | |
| Stage4 | 0.938 | 2.556 | 0.658 | 0.704 | 9.275 | 1.427 | 0.154 | |

Rsquare = 0.198 (max possible = 9.46e-01 )
Likelihood ratio test p = 4.4e-01
Wald test p = 6.18e-01
Score (logrank) test p = 4.51e-01

**ROR1 in COAD (n=458):**
Model: Surv(OS, EVENT) ~ `ROR1` + Age + Gender + Race + Stage
268 patients with 64 dying ( 190 missing obs. )

| | coef | HR | se(coef) | 95%CI_l | 95%CI_u | z | p | signif |
|---|---|---|---|---|---|---|---|---|
| ROR1 | 0.26 | 1.297 | 0.247 | 0.799 | 2.107 | 1.052 | 0.293 | |
| Age | 0.029 | 1.029 | 0.011 | 1.007 | 1.052 | 2.537 | 0.011 | * |
| Gendermale | 0.173 | 1.189 | 0.268 | 0.702 | 2.012 | 0.644 | 0.52 | |
| RaceBlack | -0.451 | 0.637 | 0.808 | 0.131 | 3.104 | -0.558 | 0.577 | |
| RaceWhite | -0.573 | 0.564 | 0.758 | 0.128 | 2.492 | -0.755 | 0.45 | |
| Stage2 | 0.195 | 1.216 | 0.562 | 0.404 | 3.66 | 0.348 | 0.728 | |
| Stage3 | 0.802 | 2.23 | 0.55 | 0.76 | 6.547 | 1.459 | 0.144 | |
| Stage4 | 1.927 | 6.871 | 0.556 | 2.309 | 20.448 | 3.464 | 0.001 | ** |

Rsquare = 0.112 (max possible = 8.99e-01 )
Likelihood ratio test p = 9.51e-05
Wald test p = 4.19e-05
Score (logrank) test p = 6.4e-06

**ROR1 in DLBC (n=48):**
Model: Surv(OS, EVENT) ~ `ROR1` + Age + Gender + Race
47 patients with 9 dying ( 1 missing obs. )

| | coef | HR | se(coef) | 95%CI_l | 95%CI_u | z | p | signif |
|---|---|---|---|---|---|---|---|---|

| | | | | | | | | |
|---|---|---|---|---|---|---|---|---|
| ROR1 | -0.403 | 0.668 | 1.289 | 0.053 | 8.367 | -0.312 | 0.755 | |
| Age | -0.002 | 0.998 | 0.031 | 0.94 | 1.06 | -0.063 | 0.95 | |
| Gendermale | 0.452 | 1.572 | 0.8 | 0.328 | 7.54 | 0.565 | 0.572 | |
| RaceBlack | 1.01 | 2.747 | 1.277 | 0.225 | 33.529 | 0.792 | 0.429 | |
| RaceWhite | -1.534 | 0.216 | 0.926 | 0.035 | 1.325 | -1.656 | 0.098 | . |

Rsquare = 0.102 (max possible = 6.72e-01 )
Likelihood ratio test p = 4.07e-01
Wald test p = 4.43e-01
Score (logrank) test p = 2.88e-01

**ROR1 in ESCA (n=185):**
Model: Surv(OS, EVENT) ~ `ROR1` + Age + Gender + Race + Stage
142 patients with 47 dying ( 43 missing obs. )

| | coef | HR | se(coef) | 95%CI_l | 95%CI_u | z | p | signif |
|---|---|---|---|---|---|---|---|---|
| ROR1 | -0.349 | 0.706 | 0.157 | 0.518 | 0.961 | -2.216 | 0.027 | * |
| Age | 0.019 | 1.019 | 0.014 | 0.991 | 1.048 | 1.315 | 0.188 | |
| Gendermale | 0.546 | 1.726 | 0.541 | 0.598 | 4.988 | 1.009 | 0.313 | |
| RaceBlack | 0.165 | 1.179 | 1.07 | 0.145 | 9.599 | 0.154 | 0.878 | |
| RaceWhite | 0.136 | 1.145 | 0.452 | 0.472 | 2.777 | 0.3 | 0.764 | |
| Stage2 | 0.802 | 2.23 | 0.651 | 0.622 | 7.988 | 1.232 | 0.218 | |
| Stage3 | 1.531 | 4.624 | 0.664 | 1.259 | 16.978 | 2.307 | 0.021 | * |
| Stage4 | 2.903 | 18.226 | 0.763 | 4.089 | 81.234 | 3.807 | 0 | *** |

Rsquare = 0.168 (max possible = 9.3e-01 )
Likelihood ratio test p = 1.02e-03
Wald test p = 4.37e-04
Score (logrank) test p = 2.56e-05

**ROR1 in GBM (n=153):**
Model: Surv(OS, EVENT) ~ `ROR1` + Age + Gender + Race
149 patients with 119 dying ( 4 missing obs. )

| | coef | HR | se(coef) | 95%CI_l | 95%CI_u | z | p | signif |
|---|---|---|---|---|---|---|---|---|
| ROR1 | -0.233 | 0.792 | 0.156 | 0.584 | 1.075 | -1.494 | 0.135 | |
| Age | 0.032 | 1.032 | 0.008 | 1.016 | 1.049 | 3.932 | 0 | *** |
| Gendermale | -0.071 | 0.932 | 0.197 | 0.633 | 1.372 | -0.357 | 0.721 | |
| RaceBlack | 0.468 | 1.597 | 0.723 | 0.387 | 6.595 | 0.647 | 0.517 | |
| RaceWhite | -0.19 | 0.827 | 0.607 | 0.251 | 2.717 | -0.314 | 0.754 | |

Rsquare = 0.124 (max possible = 9.98e-01 )
Likelihood ratio test p = 1.44e-03
Wald test p = 2.84e-03
Score (logrank) test p = 2.61e-03

**ROR1 in HNSC (n=522):**
Model: Surv(OS, EVENT) ~ `ROR1` + Age + Gender + Race + Stage
431 patients with 184 dying ( 91 missing obs. )

| | coef | HR | se(coef) | 95%CI_l | 95%CI_u | z | p | signif |
|---|---|---|---|---|---|---|---|---|
| ROR1 | 0.148 | 1.159 | 0.125 | 0.908 | 1.481 | 1.184 | 0.236 | |
| Age | 0.021 | 1.021 | 0.007 | 1.007 | 1.036 | 2.861 | 0.004 | ** |
| Gendermale | -0.271 | 0.763 | 0.166 | 0.55 | 1.057 | -1.629 | 0.103 | |
| RaceBlack | 0.069 | 1.072 | 0.559 | 0.358 | 3.209 | 0.124 | 0.901 | |
| RaceWhite | -0.323 | 0.724 | 0.513 | 0.265 | 1.978 | -0.63 | 0.529 | |
| Stage2 | 0.775 | 2.17 | 0.545 | 0.746 | 6.317 | 1.422 | 0.155 | |
| Stage3 | 0.951 | 2.588 | 0.54 | 0.899 | 7.452 | 1.763 | 0.078 | . |
| Stage4 | 1.381 | 3.981 | 0.511 | 1.461 | 10.845 | 2.702 | 0.007 | ** |

Rsquare = 0.07 (max possible = 9.9e-01 )
Likelihood ratio test p = 1.26e-04
Wald test p = 3.09e-04
Score (logrank) test p = 2.12e-04

**ROR1 in HNSC-HPV+ (n=98):**
Model: Surv(OS, EVENT) ~ `ROR1` + Age + Gender + Race + Stage
67 patients with 24 dying ( 31 missing obs. )

| | coef | HR | se(coef) | 95%CI_l | 95%CI_u | z | p | signif |
|---|---|---|---|---|---|---|---|---|
| ROR1 | 0.243 | 1.275 | 0.307 | 0.698 | 2.329 | 0.791 | 0.429 | |
| Age | 0.006 | 1.006 | 0.024 | 0.96 | 1.053 | 0.237 | 0.812 | |
| Gendermale | -0.061 | 0.941 | 0.524 | 0.337 | 2.627 | -0.117 | 0.907 | |

| | | | | | | | |
|---|---|---|---|---|---|---|---|
| RaceBlack | 18.255 | 84761125.65 | 12151.84 | 0 | Inf | 0.002 | 0.999 |
| RaceWhite | 17.353 | 34377091.71 | 12151.84 | 0 | Inf | 0.001 | 0.999 |
| Stage2 | 17.175 | 28767347.72 | 5343.627 | 0 | Inf | 0.003 | 0.997 |
| Stage3 | 16.16 | 10428062.73 | 5343.627 | 0 | Inf | 0.003 | 0.998 |
| Stage4 | 17.331 | 33646617.36 | 5343.627 | 0 | Inf | 0.003 | 0.997 |

Rsquare = 0.081 (max possible = 9.24e-01 )
Likelihood ratio test p = 6.83e-01
Wald test p = 9.35e-01
Score (logrank) test p = 8.22e-01

**ROR1 in HNSC-HPV- (n=422):**
Model: Surv(OS, EVENT) ~ `ROR1` + Age + Gender + Race + Stage
364 patients with 160 dying ( 58 missing obs. )

| | coef | HR | se(coef) | 95%CI_l | 95%CI_u | z | p | signif |
|---|---|---|---|---|---|---|---|---|
| ROR1 | 0.118 | 1.125 | 0.14 | 0.855 | 1.479 | 0.842 | 0.4 | |
| Age | 0.024 | 1.024 | 0.008 | 1.008 | 1.041 | 3.009 | 0.003 | ** |
| Gendermale | -0.294 | 0.746 | 0.176 | 0.528 | 1.053 | -1.665 | 0.096 | . |
| RaceBlack | -0.063 | 0.939 | 0.567 | 0.309 | 2.851 | -0.112 | 0.911 | |
| RaceWhite | -0.445 | 0.641 | 0.516 | 0.233 | 1.762 | -0.862 | 0.389 | |
| Stage2 | 0.55 | 1.734 | 0.555 | 0.584 | 5.148 | 0.991 | 0.322 | |
| Stage3 | 0.873 | 2.394 | 0.544 | 0.824 | 6.956 | 1.604 | 0.109 | |
| Stage4 | 1.29 | 3.634 | 0.514 | 1.328 | 9.944 | 2.512 | 0.012 | * |

Rsquare = 0.081 (max possible = 9.89e-01 )
Likelihood ratio test p = 1.57e-04
Wald test p = 3.74e-04
Score (logrank) test p = 2.72e-04

**ROR1 in KICH (n=66):**
Model: Surv(OS, EVENT) ~ `ROR1` + Age + Gender + Race + Stage
63 patients with 9 dying ( 3 missing obs. )

| | coef | HR | se(coef) | 95%CI_l | 95%CI_u | z | p | signif |
|---|---|---|---|---|---|---|---|---|
| ROR1 | 0.127 | 1.14E+00 | 0.51 | 0.418 | 3.08E+00 | 0.249 | 0.804 | |
| Age | 0.072 | 1.07E+00 | 0.029 | 1.016 | 1.14E+00 | 2.499 | 0.012 | * |
| Gendermale | -0.885 | 4.13E-01 | 0.728 | 0.099 | 1.72E+00 | -1.215 | 0.224 | |
| RaceBlack | -17.003 | 0.00E+00 | 6326.393 | 0 | Inf | -0.003 | 0.998 | |
| RaceWhite | -1.837 | 1.59E-01 | 1.161 | 0.016 | 1.55E+00 | -1.582 | 0.114 | |
| Stage2 | 16.083 | 9.65E+06 | 0.849 | 1826607.177 | 5.10E+07 | 18.934 | 0 | **** |
| Stage3 | 17.145 | 2.79E+07 | 0.776 | 6102805.105 | 1.28E+08 | 22.097 | 0 | **** |
| Stage4 | 19.591 | 3.22E+08 | 0.899 | 55280881.16 | 1.88E+09 | 21.78 | 0 | **** |

Rsquare = 0.345 (max possible = 6.71e-01 )
Likelihood ratio test p = 7.96e-04
Wald test p = 3.7e-282
Score (logrank) test p = 3.63e-09

**ROR1 in KIRC (n=533):**
Model: Surv(OS, EVENT) ~ `ROR1` + Age + Gender + Race + Stage
519 patients with 170 dying ( 14 missing obs. )

| | coef | HR | se(coef) | 95%CI_l | 95%CI_u | z | p | signif |
|---|---|---|---|---|---|---|---|---|
| ROR1 | -0.447 | 0.64 | 0.116 | 0.509 | 0.804 | -3.836 | 0 | *** |
| Age | 0.031 | 1.032 | 0.007 | 1.017 | 1.047 | 4.206 | 0 | *** |
| Gendermale | -0.144 | 0.866 | 0.165 | 0.627 | 1.197 | -0.87 | 0.384 | |
| RaceBlack | 0.372 | 1.451 | 1.053 | 0.184 | 11.428 | 0.353 | 0.724 | |
| RaceWhite | 0.551 | 1.735 | 1.013 | 0.238 | 12.642 | 0.544 | 0.587 | |
| Stage2 | 0.232 | 1.262 | 0.321 | 0.672 | 2.368 | 0.723 | 0.47 | |
| Stage3 | 0.91 | 2.484 | 0.213 | 1.635 | 3.773 | 4.265 | 0 | *** |
| Stage4 | 1.884 | 6.579 | 0.201 | 4.439 | 9.749 | 9.386 | 0 | *** |

Rsquare = 0.236 (max possible = 9.75e-01 )
Likelihood ratio test p = 2.78e-26
Wald test p = 5.31e-26
Score (logrank) test p = 4.89e-32

**ROR1 in KIRP (n=290):**
Model: Surv(OS, EVENT) ~ `ROR1` + Age + Gender + Race + Stage
243 patients with 38 dying ( 47 missing obs. )

| | coef | HR | se(coef) | 95%CI_l | 95%CI_u | z | p | signif |
|---|---|---|---|---|---|---|---|---|

| | | | | | | | | |
|---|---|---|---|---|---|---|---|---|
| ROR1 | 0.172 | 1.187 | 0.181 | 0.832 | 1.694 | 0.948 | 0.343 | |
| Age | 0.014 | 1.014 | 0.016 | 0.983 | 1.045 | 0.871 | 0.384 | |
| Gendermale | -0.253 | 0.777 | 0.364 | 0.38 | 1.586 | -0.693 | 0.488 | |
| RaceBlack | -2.08 | 0.125 | 1.158 | 0.013 | 1.209 | -1.796 | 0.073 | . |
| RaceWhite | -2.299 | 0.1 | 1.149 | 0.011 | 0.955 | -2 | 0.045 | * |
| Stage2 | 0.001 | 1.001 | 0.769 | 0.222 | 4.517 | 0.001 | 0.999 | |
| Stage3 | 1.428 | 4.17 | 0.389 | 1.946 | 8.939 | 3.671 | 0 | *** |
| Stage4 | 2.703 | 14.92 | 0.464 | 6.014 | 37.014 | 5.83 | 0 | *** |

Rsquare = 0.15 (max possible = 7.65e-01 )
Likelihood ratio test p = 4.22e-06
Wald test p = 5.48e-07
Score (logrank) test p = 3.47e-11

**ROR1 in LAML (n=173):**
Model: Surv(OS, EVENT) ~ `ROR1` + Age + Gender + Race
149 patients with 93 dying ( 24 missing obs. )

| | coef | HR | se(coef) | 95%CI_l | 95%CI_u | z | p | signif |
|---|---|---|---|---|---|---|---|---|
| ROR1 | 0.536 | 1.709 | 0.543 | 0.59 | 4.949 | 0.987 | 0.323 | |
| Age | 0.036 | 1.037 | 0.008 | 1.02 | 1.054 | 4.357 | 0 | *** |
| Gendermale | -0.115 | 0.891 | 0.213 | 0.587 | 1.352 | -0.542 | 0.588 | |
| RaceBlack | -0.418 | 0.659 | 1.108 | 0.075 | 5.781 | -0.377 | 0.706 | |
| RaceWhite | -0.785 | 0.456 | 1.021 | 0.062 | 3.374 | -0.769 | 0.442 | |

Rsquare = 0.161 (max possible = 9.96e-01 )
Likelihood ratio test p = 8.5e-05
Wald test p = 2.12e-04
Score (logrank) test p = 1.28e-04

**ROR1 in LGG (n=516):**
Model: Surv(OS, EVENT) ~ `ROR1` + Age + Gender + Race
500 patients with 124 dying ( 16 missing obs. )

| | coef | HR | se(coef) | 95%CI_l | 95%CI_u | z | p | signif |
|---|---|---|---|---|---|---|---|---|
| ROR1 | 0.933 | 2.543 | 0.133 | 1.961 | 3.297 | 7.042 | 0 | *** |
| Age | 0.059 | 1.061 | 0.007 | 1.046 | 1.076 | 8.146 | 0 | *** |
| Gendermale | 0.241 | 1.273 | 0.191 | 0.876 | 1.85 | 1.266 | 0.206 | |
| RaceBlack | 15.336 | 4573467.195 | 2533.119 | 0 | Inf | 0.006 | 0.995 | |
| RaceWhite | 15.778 | 7117335.446 | 2533.119 | 0 | Inf | 0.006 | 0.995 | |

Rsquare = 0.188 (max possible = 9.16e-01 )
Likelihood ratio test p = 6.95e-21
Wald test p = 1.24e-21
Score (logrank) test p = 2.4e-25

**ROR1 in LIHC (n=371):**
Model: Surv(OS, EVENT) ~ `ROR1` + Age + Gender + Race + Stage
330 patients with 112 dying ( 41 missing obs. )

| | coef | HR | se(coef) | 95%CI_l | 95%CI_u | z | p | signif |
|---|---|---|---|---|---|---|---|---|
| ROR1 | 0.295 | 1.343 | 0.158 | 0.985 | 1.831 | 1.862 | 0.063 | . |
| Age | 0.012 | 1.012 | 0.008 | 0.997 | 1.028 | 1.557 | 0.119 | |
| Gendermale | -0.174 | 0.84 | 0.214 | 0.552 | 1.279 | -0.812 | 0.417 | |
| RaceBlack | 0.668 | 1.95 | 0.481 | 0.76 | 5.001 | 1.39 | 0.165 | |
| RaceWhite | -0.103 | 0.902 | 0.224 | 0.582 | 1.399 | -0.461 | 0.645 | |
| Stage2 | 0.263 | 1.3 | 0.257 | 0.786 | 2.151 | 1.023 | 0.306 | |
| Stage3 | 0.974 | 2.648 | 0.224 | 1.706 | 4.109 | 4.342 | 0 | *** |
| Stage4 | 1.734 | 5.664 | 0.616 | 1.693 | 18.951 | 2.814 | 0.005 | ** |

Rsquare = 0.087 (max possible = 9.67e-01 )
Likelihood ratio test p = 2.07e-04
Wald test p = 9.67e-05
Score (logrank) test p = 3.12e-05

**ROR1 in LUAD (n=515):**
Model: Surv(OS, EVENT) ~ `ROR1` + Age + Gender + Race + Stage
439 patients with 160 dying ( 76 missing obs. )

| | coef | HR | se(coef) | 95%CI_l | 95%CI_u | z | p | signif |
|---|---|---|---|---|---|---|---|---|
| ROR1 | -0.029 | 0.972 | 0.093 | 0.811 | 1.165 | -0.311 | 0.756 | |
| Age | 0.005 | 1.005 | 0.009 | 0.988 | 1.022 | 0.594 | 0.552 | |
| Gendermale | 0.041 | 1.042 | 0.163 | 0.757 | 1.434 | 0.254 | 0.8 | |

| | | | | | | | | |
|---|---|---|---|---|---|---|---|---|
| RaceBlack | 0.936 | 2.55 | 1.036 | 0.335 | 19.429 | 0.904 | 0.366 | |
| RaceWhite | 1.162 | 3.196 | 1.007 | 0.444 | 23.017 | 1.154 | 0.249 | |
| Stage2 | 0.85 | 2.34 | 0.195 | 1.595 | 3.432 | 4.349 | 0 | *** |
| Stage3 | 1.074 | 2.927 | 0.209 | 1.943 | 4.41 | 5.136 | 0 | *** |
| Stage4 | 1.149 | 3.154 | 0.317 | 1.695 | 5.868 | 3.626 | 0 | *** |

Rsquare = 0.089 (max possible = 9.77e-01 )
Likelihood ratio test p = 2.16e-06
Wald test p = 3.87e-06
Score (logrank) test p = 9.72e-07

**ROR1 in LUSC (n=501):**
Model: Surv(OS, EVENT) ~ `ROR1` + Age + Gender + Race + Stage
382 patients with 168 dying ( 119 missing obs. )

| | coef | HR | se(coef) | 95%CI_l | 95%CI_u | z | p | signif |
|---|---|---|---|---|---|---|---|---|
| ROR1 | 0.171 | 1.186 | 0.105 | 0.966 | 1.458 | 1.627 | 0.104 | |
| Age | 0.015 | 1.016 | 0.009 | 0.998 | 1.034 | 1.697 | 0.09 | . |
| Gendermale | 0.472 | 1.604 | 0.191 | 1.104 | 2.33 | 2.477 | 0.013 | * |
| RaceBlack | 0.218 | 1.243 | 0.619 | 0.37 | 4.179 | 0.352 | 0.725 | |
| RaceWhite | -0.332 | 0.718 | 0.571 | 0.235 | 2.195 | -0.582 | 0.561 | |
| Stage2 | 0.236 | 1.266 | 0.182 | 0.885 | 1.81 | 1.291 | 0.197 | |
| Stage3 | 0.606 | 1.832 | 0.212 | 1.209 | 2.778 | 2.853 | 0.004 | ** |
| Stage4 | 1.199 | 3.318 | 0.664 | 0.903 | 12.187 | 1.807 | 0.071 | . |

Rsquare = 0.058 (max possible = 9.88e-01 )
Likelihood ratio test p = 3.36e-03
Wald test p = 1.89e-03
Score (logrank) test p = 1.34e-03

**ROR1 in MESO (n=87):**
Model: Surv(OS, EVENT) ~ `ROR1` + Age + Gender + Race + Stage
86 patients with 73 dying ( 1 missing obs. )

| | coef | HR | se(coef) | 95%CI_l | 95%CI_u | z | p | signif |
|---|---|---|---|---|---|---|---|---|
| ROR1 | 0.235 | 1.265 | 0.119 | 1.002 | 1.596 | 1.981 | 0.048 | * |
| Age | 0.018 | 1.019 | 0.016 | 0.988 | 1.05 | 1.178 | 0.239 | |
| Gendermale | -0.263 | 0.769 | 0.335 | 0.399 | 1.483 | -0.784 | 0.433 | |
| RaceBlack | -0.791 | 0.453 | 1.598 | 0.02 | 10.39 | -0.495 | 0.62 | |
| RaceWhite | -1.059 | 0.347 | 1.074 | 0.042 | 2.845 | -0.986 | 0.324 | |
| Stage2 | -0.226 | 0.798 | 0.458 | 0.325 | 1.959 | -0.492 | 0.622 | |
| Stage3 | -0.04 | 0.961 | 0.409 | 0.431 | 2.143 | -0.097 | 0.922 | |
| Stage4 | -0.228 | 0.796 | 0.459 | 0.324 | 1.957 | -0.496 | 0.62 | |

Rsquare = 0.084 (max possible = 9.98e-01 )
Likelihood ratio test p = 4.78e-01
Wald test p = 4.93e-01
Score (logrank) test p = 4.8e-01

**ROR1 in OV (n=303):**
Model: Surv(OS, EVENT) ~ `ROR1` + Age + Race
289 patients with 178 dying ( 14 missing obs. )

| | coef | HR | se(coef) | 95%CI_l | 95%CI_u | z | p | signif |
|---|---|---|---|---|---|---|---|---|
| ROR1 | 0.011 | 1.011 | 0.081 | 0.862 | 1.186 | 0.137 | 0.891 | |
| Age | 0.031 | 1.032 | 0.008 | 1.017 | 1.047 | 4.147 | 0 | *** |
| RaceBlack | 0.112 | 1.118 | 0.564 | 0.37 | 3.381 | 0.198 | 0.843 | |
| RaceWhite | -0.068 | 0.934 | 0.515 | 0.34 | 2.564 | -0.133 | 0.894 | |

Rsquare = 0.059 (max possible = 9.97e-01 )
Likelihood ratio test p = 1.5e-03
Wald test p = 1.34e-03
Score (logrank) test p = 1.2e-03

**ROR1 in PAAD (n=179):**
Model: Surv(OS, EVENT) ~ `ROR1` + Age + Gender + Race + Stage
171 patients with 90 dying ( 8 missing obs. )

| | coef | HR | se(coef) | 95%CI_l | 95%CI_u | z | p | signif |
|---|---|---|---|---|---|---|---|---|
| ROR1 | 0.213 | 1.238 | 0.142 | 0.938 | 1.634 | 1.505 | 0.132 | |
| Age | 0.023 | 1.023 | 0.011 | 1.001 | 1.046 | 2.089 | 0.037 | * |
| Gendermale | -0.246 | 0.782 | 0.215 | 0.514 | 1.191 | -1.145 | 0.252 | |
| RaceBlack | -0.022 | 0.978 | 0.701 | 0.248 | 3.865 | -0.032 | 0.975 | |

| | | | | | | | | |
|---|---|---|---|---|---|---|---|---|
| RaceWhite | 0.269 | 1.308 | 0.479 | 0.511 | 3.347 | 0.56 | 0.575 | |
| Stage2 | 0.846 | 2.33 | 0.439 | 0.986 | 5.505 | 1.928 | 0.054 | . |
| Stage3 | 0.151 | 1.163 | 1.088 | 0.138 | 9.806 | 0.139 | 0.889 | |
| Stage4 | 0.286 | 1.331 | 0.834 | 0.259 | 6.825 | 0.342 | 0.732 | |

Rsquare = 0.087 (max possible = 9.9e-01 )
Likelihood ratio test p = 5.05e-02
Wald test p = 8.26e-02
Score (logrank) test p = 6.92e-02

**ROR1 in PCPG (n=181):**
Model: Surv(OS, EVENT) ~ `ROR1` + Age + Gender + Race
176 patients with 7 dying ( 5 missing obs. )

| | coef | HR | se(coef) | 95%CI_l | 95%CI_u | z | p | signif |
|---|---|---|---|---|---|---|---|---|
| ROR1 | 0.294 | 1.342 | 0.957 | 0.206 | 8.763 | 0.308 | 0.758 | |
| Age | 0.031 | 1.031 | 0.025 | 0.982 | 1.083 | 1.236 | 0.216 | |
| Gendermale | 1.552 | 4.719 | 0.954 | 0.727 | 30.627 | 1.626 | 0.104 | |
| RaceBlack | 0.128 | 1.137 | 21211.75 | 0 | Inf | 0 | 1 | <NA> |
| RaceWhite | 17.777 | 52532118.43 | 17295.11 | 0 | Inf | 0.001 | 0.999 | |

Rsquare = 0.032 (max possible = 2.95e-01 )
Likelihood ratio test p = 3.38e-01
Wald test p = 7.28e-01
Score (logrank) test p = 4.26e-01

**ROR1 in PRAD (n=498):**
Model: Surv(OS, EVENT) ~ `ROR1` + Age + Race
481 patients with 10 dying ( 17 missing obs. )

| | coef | HR | se(coef) | 95%CI_l | 95%CI_u | z | p | signif |
|---|---|---|---|---|---|---|---|---|
| ROR1 | -0.496 | 0.609 | 0.463 | 0.246 | 1.508 | -1.073 | 0.283 | |
| Age | 0.044 | 1.045 | 0.052 | 0.944 | 1.157 | 0.844 | 0.399 | |
| RaceBlack | 15.809 | 7343392.664 | 6678.002 | 0 | Inf | 0.002 | 0.998 | |
| RaceWhite | 16.195 | 10797687.38 | 6678.002 | 0 | Inf | 0.002 | 0.998 | |

Rsquare = 0.006 (max possible = 1.72e-01 )
Likelihood ratio test p = 6.01e-01
Wald test p = 7.1e-01
Score (logrank) test p = 6.64e-01

**ROR1 in READ (n=166):**
Model: Surv(OS, EVENT) ~ `ROR1` + Age + Gender + Race + Stage
78 patients with 14 dying ( 88 missing obs. )

| | coef | HR | se(coef) | 95%CI_l | 95%CI_u | z | p | signif |
|---|---|---|---|---|---|---|---|---|
| ROR1 | 1.212 | 3.361 | 0.664 | 0.916 | 12.339 | 1.827 | 0.068 | . |
| Age | 0.141 | 1.151 | 0.053 | 1.038 | 1.277 | 2.663 | 0.008 | ** |
| Gendermale | -0.009 | 0.991 | 0.701 | 0.251 | 3.919 | -0.013 | 0.99 | |
| RaceBlack | 12.85 | 380660.485 | 6979.641 | 0 | Inf | 0.002 | 0.999 | |
| RaceWhite | 11.073 | 64436.966 | 6979.641 | 0 | Inf | 0.002 | 0.999 | |
| Stage2 | -1.765 | 0.171 | 1.261 | 0.014 | 2.028 | -1.399 | 0.162 | |
| Stage3 | -0.215 | 0.806 | 0.913 | 0.135 | 4.828 | -0.236 | 0.814 | |
| Stage4 | -0.462 | 0.63 | 1.014 | 0.086 | 4.597 | -0.455 | 0.649 | |

Rsquare = 0.236 (max possible = 7.16e-01 )
Likelihood ratio test p = 7.24e-03
Wald test p = 1e-01
Score (logrank) test p = 1.66e-02

**ROR1 in SARC (n=260):**
Model: Surv(OS, EVENT) ~ `ROR1` + Age + Gender + Race
248 patients with 94 dying ( 12 missing obs. )

| | coef | HR | se(coef) | 95%CI_l | 95%CI_u | z | p | signif |
|---|---|---|---|---|---|---|---|---|
| ROR1 | 0.118 | 1.125 | 0.083 | 0.957 | 1.323 | 1.423 | 0.155 | |
| Age | 0.021 | 1.021 | 0.008 | 1.005 | 1.038 | 2.602 | 0.009 | ** |
| Gendermale | -0.115 | 0.892 | 0.217 | 0.583 | 1.363 | -0.529 | 0.597 | |
| RaceBlack | -0.019 | 0.981 | 1.074 | 0.119 | 8.059 | -0.018 | 0.986 | |
| RaceWhite | -0.368 | 0.692 | 1.019 | 0.094 | 5.101 | -0.361 | 0.718 | |

Rsquare = 0.04 (max possible = 9.74e-01 )
Likelihood ratio test p = 6.86e-02
Wald test p = 8.44e-02

Score (logrank) test p = 8.46e-02

**ROR1 in SKCM (n=471):**
Model: Surv(OS, EVENT) ~ `ROR1` + Age + Gender + Race + Stage
400 patients with 191 dying ( 71 missing obs. )

| | coef | HR | se(coef) | 95%CI_l | 95%CI_u | z | p | signif |
|---|---|---|---|---|---|---|---|---|
| ROR1 | -0.01 | 0.99 | 0.085 | 0.838 | 1.169 | -0.123 | 0.902 | |
| Age | 0.02 | 1.02 | 0.005 | 1.01 | 1.03 | 3.882 | 0 | *** |
| Gendermale | -0.02 | 0.981 | 0.156 | 0.722 | 1.332 | -0.125 | 0.901 | |
| RaceWhite | -1.359 | 0.257 | 0.401 | 0.117 | 0.564 | -3.387 | 0.001 | ** |
| Stage2 | 0.26 | 1.297 | 0.216 | 0.849 | 1.983 | 1.204 | 0.229 | |
| Stage3 | 0.588 | 1.8 | 0.202 | 1.212 | 2.674 | 2.914 | 0.004 | ** |
| Stage4 | 1.235 | 3.437 | 0.349 | 1.735 | 6.808 | 3.54 | 0 | *** |

Rsquare = 0.101 (max possible = 9.92e-01 )
Likelihood ratio test p = 4.06e-07
Wald test p = 1.05e-07
Score (logrank) test p = 1.78e-08

**ROR1 in SKCM-Primary (n=103):**
Model: Surv(OS, EVENT) ~ `ROR1` + Age + Gender + Race + Stage
94 patients with 26 dying ( 9 missing obs. )

| | coef | HR | se(coef) | 95%CI_l | 95%CI_u | z | p | signif |
|---|---|---|---|---|---|---|---|---|
| ROR1 | 0.009 | 1.01E+00 | 0.267 | 0.598 | 1.701 | 0.032 | 0.974 | |
| Age | 0.013 | 1.01E+00 | 0.016 | 0.982 | 1.045 | 0.794 | 0.427 | |
| Gendermale | 0.235 | 1.27E+00 | 0.43 | 0.545 | 2.937 | 0.548 | 0.584 | |
| RaceWhite | -1.293 | 2.74E-01 | 0.615 | 0.082 | 0.916 | -2.102 | 0.036 | * |
| Stage2 | 17.475 | 3.88E+07 | 6203.221 | 0 | Inf | 0.003 | 0.998 | |
| Stage3 | 17.967 | 6.35E+07 | 6203.221 | 0 | Inf | 0.003 | 0.998 | |
| Stage4 | 20.127 | 5.51E+08 | 6203.221 | 0 | Inf | 0.003 | 0.997 | |

Rsquare = 0.146 (max possible = 8.69e-01 )
Likelihood ratio test p = 3.86e-02
Wald test p = 3.47e-02
Score (logrank) test p = 2.44e-03

**ROR1 in SKCM-Metastasis (n=368):**
Model: Surv(OS, EVENT) ~ `ROR1` + Age + Gender + Race + Stage
306 patients with 165 dying ( 62 missing obs. )

| | coef | HR | se(coef) | 95%CI_l | 95%CI_u | z | p | signif |
|---|---|---|---|---|---|---|---|---|
| ROR1 | 0.018 | 1.018 | 0.09 | 0.853 | 1.215 | 0.196 | 0.845 | |
| Age | 0.022 | 1.023 | 0.006 | 1.011 | 1.034 | 4.017 | 0 | *** |
| Gendermale | -0.03 | 0.97 | 0.171 | 0.694 | 1.355 | -0.178 | 0.859 | |
| RaceWhite | -1.16 | 0.313 | 0.599 | 0.097 | 1.013 | -1.938 | 0.053 | . |
| Stage2 | 0.144 | 1.155 | 0.227 | 0.739 | 1.803 | 0.632 | 0.527 | |
| Stage3 | 0.54 | 1.716 | 0.206 | 1.146 | 2.57 | 2.622 | 0.009 | ** |
| Stage4 | 0.971 | 2.641 | 0.396 | 1.215 | 5.738 | 2.453 | 0.014 | * |

Rsquare = 0.103 (max possible = 9.95e-01 )
Likelihood ratio test p = 2.48e-05
Wald test p = 1.97e-05
Score (logrank) test p = 9.79e-06

**ROR1 in STAD (n=415):**
Model: Surv(OS, EVENT) ~ `ROR1` + Age + Gender + Race + Stage
323 patients with 125 dying ( 92 missing obs. )

| | coef | HR | se(coef) | 95%CI_l | 95%CI_u | z | p | signif |
|---|---|---|---|---|---|---|---|---|
| ROR1 | 0.233 | 1.263 | 0.088 | 1.063 | 1.5 | 2.661 | 0.008 | ** |
| Age | 0.033 | 1.034 | 0.01 | 1.014 | 1.054 | 3.403 | 0.001 | ** |
| Gendermale | 0.108 | 1.114 | 0.192 | 0.765 | 1.622 | 0.563 | 0.574 | |
| RaceBlack | 0.226 | 1.254 | 0.441 | 0.528 | 2.974 | 0.513 | 0.608 | |
| RaceWhite | -0.033 | 0.968 | 0.236 | 0.609 | 1.537 | -0.14 | 0.889 | |
| Stage2 | 0.594 | 1.811 | 0.383 | 0.855 | 3.835 | 1.551 | 0.121 | |
| Stage3 | 1.025 | 2.788 | 0.359 | 1.381 | 5.63 | 2.859 | 0.004 | ** |
| Stage4 | 1.838 | 6.284 | 0.486 | 2.426 | 16.273 | 3.786 | 0 | *** |

Rsquare = 0.085 (max possible = 9.81e-01 )
Likelihood ratio test p = 3.66e-04
Wald test p = 4.94e-04

Score (logrank) test p = 4e-04

**ROR1 in TGCT (n=150):**
Model: Surv(OS, EVENT) ~ `ROR1` + Age + Race + Stage
77 patients with 2 dying ( 73 missing obs. )

| | coef | HR | se(coef) | 95%CI_l | 95%CI_u | z | p | signif |
|---|---|---|---|---|---|---|---|---|
| ROR1 | -7.35 | 0.001 | 12310.66 | 0 | Inf | -0.001 | 1 | <NA> |
| Age | -2.392 | 0.091 | 1734.582 | 0 | Inf | -0.001 | 0.999 | |
| RaceBlack | 5.344 | 209.278 | 3804039 | 0 | Inf | 0 | 1 | <NA> |
| RaceWhite | -38.366 | 0 | 3811962 | 0 | Inf | 0 | 1 | <NA> |
| Stage2 | 1.531 | 4.624 | 33496.55 | 0 | Inf | 0 | 1 | <NA> |
| Stage3 | 11.142 | 69011.398 | 103956.3 | 0 | Inf | 0 | 1 | <NA> |

Rsquare = 0.137 (max possible = 1.37e-01 )
Likelihood ratio test p = 7.73e-02
Wald test p = 1e+00
Score (logrank) test p = 8.82e-03

**ROR1 in THCA (n=509):**
Model: Surv(OS, EVENT) ~ `ROR1` + Age + Gender + Race + Stage
411 patients with 16 dying ( 98 missing obs. )

| | coef | HR | se(coef) | 95%CI_l | 95%CI_u | z | p | signif |
|---|---|---|---|---|---|---|---|---|
| ROR1 | 0.381 | 1.46E+00 | 0.463 | 0.59 | 3.628 | 0.821 | 0.411 | |
| Age | 0.153 | 1.17E+00 | 0.03 | 1.099 | 1.235 | 5.137 | 0 | *** |
| Gendermale | 0.442 | 1.56E+00 | 0.584 | 0.495 | 4.89 | 0.756 | 0.45 | |
| RaceBlack | 18.545 | 1.13E+08 | 8109.587 | 0 | Inf | 0.002 | 0.998 | |
| RaceWhite | 18.17 | 7.78E+07 | 8109.587 | 0 | Inf | 0.002 | 0.998 | |
| Stage2 | 0.322 | 1.38E+00 | 1.063 | 0.172 | 11.078 | 0.303 | 0.762 | |
| Stage3 | -0.115 | 8.92E-01 | 0.849 | 0.169 | 4.709 | -0.135 | 0.893 | |
| Stage4 | 0.715 | 2.05E+00 | 0.888 | 0.359 | 11.656 | 0.805 | 0.421 | |

Rsquare = 0.137 (max possible = 3.38e-01 )
Likelihood ratio test p = 3.53e-10
Wald test p = 2.59e-04
Score (logrank) test p = 1.53e-10

**ROR1 in THYM (n=120):**
Model: Surv(OS, EVENT) ~ `ROR1` + Age + Gender + Race
117 patients with 9 dying ( 3 missing obs. )

| | coef | HR | se(coef) | 95%CI_l | 95%CI_u | z | p | signif |
|---|---|---|---|---|---|---|---|---|
| ROR1 | -0.316 | 0.729 | 0.464 | 0.293 | 1.813 | -0.679 | 0.497 | |
| Age | 0.064 | 1.066 | 0.033 | 1 | 1.137 | 1.952 | 0.051 | . |
| Gendermale | -0.295 | 0.745 | 0.718 | 0.182 | 3.044 | -0.41 | 0.682 | |
| RaceBlack | -16.815 | 0 | 9219.931 | 0 | Inf | -0.002 | 0.999 | |
| RaceWhite | 0.489 | 1.631 | 1.1 | 0.189 | 14.088 | 0.444 | 0.657 | |

Rsquare = 0.048 (max possible = 4.44e-01 )
Likelihood ratio test p = 3.25e-01
Wald test p = 4.83e-01
Score (logrank) test p = 3.98e-01

**ROR1 in UCEC (n=545):**
Model: Surv(OS, EVENT) ~ `ROR1` + Age + Race
497 patients with 85 dying ( 48 missing obs. )

| | coef | HR | se(coef) | 95%CI_l | 95%CI_u | z | p | signif |
|---|---|---|---|---|---|---|---|---|
| ROR1 | -0.145 | 0.865 | 0.148 | 0.647 | 1.156 | -0.98 | 0.327 | |
| Age | 0.041 | 1.042 | 0.011 | 1.019 | 1.066 | 3.605 | 0 | *** |
| RaceBlack | 0.823 | 2.278 | 0.75 | 0.524 | 9.912 | 1.098 | 0.272 | |
| RaceWhite | 0.726 | 2.067 | 0.726 | 0.498 | 8.57 | 1 | 0.317 | |

Rsquare = 0.037 (max possible = 8.48e-01 )
Likelihood ratio test p = 9.47e-04
Wald test p = 1.82e-03
Score (logrank) test p = 1.64e-03

**ROR1 in UCS (n=57):**
Model: Surv(OS, EVENT) ~ `ROR1` + Age + Race
54 patients with 34 dying ( 3 missing obs. )

| | coef | HR | se(coef) | 95%CI_l | 95%CI_u | z | p | signif |
|---|---|---|---|---|---|---|---|---|

| | | | | | | | | |
|---|---|---|---|---|---|---|---|---|
| ROR1 | -0.324 | 0.723 | 0.215 | 0.475 | 1.102 | -1.509 | 0.131 | |
| Age | 0.062 | 1.064 | 0.025 | 1.013 | 1.117 | 2.47 | 0.014 | * |
| RaceBlack | 16.887 | 21567091.11 | 4076.792 | 0 | Inf | 0.004 | 0.997 | |
| RaceWhite | 17.189 | 29171120.97 | 4076.792 | 0 | Inf | 0.004 | 0.997 | |

Rsquare = 0.153 (max possible = 9.84e-01 )
Likelihood ratio test p = 6.22e-02
Wald test p = 1.23e-01
Score (logrank) test p = 8.24e-02

**ROR1 in UVM (n=80):**
Model: Surv(OS, EVENT) ~ `ROR1` + Age + Gender + Stage
79 patients with 22 dying ( 1 missing obs. )

| | coef | HR | se(coef) | 95%CI_l | 95%CI_u | z | p | signif |
|---|---|---|---|---|---|---|---|---|
| ROR1 | 1.516 | 4.553 | 1.085 | 0.543 | 38.183 | 1.397 | 0.162 | |
| Age | 0.032 | 1.033 | 0.019 | 0.995 | 1.072 | 1.7 | 0.089 | . |
| Gendermale | 0.365 | 1.44 | 0.48 | 0.562 | 3.689 | 0.76 | 0.447 | |
| Stage3 | -0.083 | 0.92 | 0.507 | 0.341 | 2.486 | -0.164 | 0.87 | |
| Stage4 | 3.792 | 44.365 | 1.225 | 4.021 | 489.522 | 3.096 | 0.002 | ** |

Rsquare = 0.238 (max possible = 8.67e-01 )
Likelihood ratio test p = 6.55e-04
Wald test p = 1.29e-03
Score (logrank) test p = 3.58e-10
